# Targeted Tuberculosis (TB) Vaccination Strategies in the United States: A Modeling Study

**DOI:** 10.64898/2026.05.11.26352914

**Authors:** Jessica E. Rothman, Kenneth G. Castro, Benjamin A. Lopman, Neel R. Gandhi, Kristin N. Nelson

## Abstract

**Background:** Tuberculosis (TB) incidence in the United States has remained elevated above pre-pandemic levels since 2021, with over 85% of cases resulting from reactivation of *Mycobacterium tuberculosis* (*Mtb*) infection. New vaccines that would prevent TB in adults are under development, but the potential health impact of a program prioritizing non-U.S.-born persons and persons with medical comorbidities, including persons living with HIV (PLWH), has not been evaluated.

**Methods:** We developed a deterministic compartmental transmission model that simulates *Mtb* infection, transmission, and progression to TB in the U.S., both in the general population and in key high-risk groups. We calibrated the model to 2024 U.S. TB surveillance data and estimated annual cases prevented, percent reduction in annual TB cases, and number needed to vaccinate (NNV, a measure of vaccine program efficiency) at equilibrium conditions for targeted vaccination strategies under optimistic and plausible scenarios, varying assumptions of vaccine efficacy, duration of protection, and achieved vaccination coverage in high-risk groups.

**Findings:** Under an optimistic scenario, vaccinating PLWH, non-U.S.-born persons, and persons with medical comorbidities (all high-risk groups) prevented 5,385 cases per year (51·8% reduction, NNV = 366). Under a more conservative plausible scenario, the same strategy prevented 1,348 cases per year (13·0% reduction, NNV = 510). The efficiency and impact of targeting strategies we considered were preserved across all sensitivity and uncertainty analyses.

**Interpretation:** Targeted vaccination of persons with *Mtb* infection in population subgroups recognized to be at high-risk for TB can reduce incidence substantially. Strategies that include non-U.S.-born persons and PLWH are most efficient and impactful.

**Funding:** American Lung Association, U.S. National Institutes of Health, and the Ferguson Fellowship.

## Introduction

Tuberculosis (TB) incidence in the United States (U.S.) has remained elevated above pre-pandemic levels since 2021, with 10,388 cases reported in 2024 at a rate of approximately 3·1 cases per 100,000 persons.^1^ As a low TB burden country, over 85% of TB cases in the U.S. result from reactivation of *Mycobacterium tuberculosis* (*Mtb*) infection acquired more than two years before disease onset, rather than from recent transmission.^2,3^ The burden falls disproportionately on non-U.S.-born persons, who account for approximately 77% of cases, most of whom acquired *Mtb* infection in higher TB-burden countries before immigrating to the U.S.^1^ Individuals with immuno-compromising medical comorbidities, including persons living with HIV (PLWH), end-stage renal disease (ESRD), and diabetes, face significantly elevated risk of progression from *Mtb* infection to TB disease.^4,5^ Tuberculosis preventive treatment (TPT) is recommended for persons at high risk of progression but uptake and completion are limited.^6^ Vaccines that prevent progression from *Mtb* infection to TB disease could complement TPT and reduce TB incidence in high-risk populations.

The Bacille Calmette-Guérin (BCG) vaccine is administered at birth in high-burden countries but provides limited protection against pulmonary TB in adults and is not currently recommended in countries with lower TB burdens such as the U.S.^6^ New vaccines for adolescents and adults have now advanced to late-stage clinical trials.^6,7^ One candidate, M72/AS01_E_, demonstrated 49·7% efficacy against progression to TB disease among adults with *Mtb* infection in Kenya, South Africa, and Zambia in a Phase IIb trial with three years of follow-up.^8^ Phase III trials are underway, and other promising candidates are in development.^9^ Vaccines that prevent progression from *Mtb* infection to TB disease, known as prevention of disease (POD) vaccines, are particularly relevant for the U.S., where the majority of TB cases result from reactivation of longstanding *Mtb* infection.^2^ If a new TB vaccine is licensed, deployment to high-burden settings will be prioritized. However, an efficacious vaccine could also be considered for use in the U.S. and other low-burden settings. In these settings, a key policy question will be which population subgroups should be prioritized for vaccination.^10^

Modeling studies suggest that adolescent and adult TB vaccination in the general population could have substantial population health impact in settings where TB incidence is high.^11–13^ The potential impact of a POD vaccine in a low-incidence setting like the U.S., where vaccination would likely be targeted toward high-risk populations, has not been evaluated. Previous models have examined broad pathways to TB elimination in the U.S. using combined interventions^14^ and the impact of age-targeted adult vaccination,^15^ but none have evaluated targeted vaccination of persons with *Mtb* infection across the risk groups that drive U.S. TB epidemiology – PLWH, persons with medical comorbidities that increase TB progression risk, and non-U.S.-born persons – with relevance for a specific vaccine candidate with the potential to be licensed. TB risk varies widely across high-risk groups in the U.S., so the choice of targeting strategy matters for both population impact and programmatic efficiency.

In this analysis, we developed a deterministic compartmental transmission model stratified by four clinical and demographic populations and calibrated to 2024 U.S. TB surveillance data.^1^ We evaluated multiple vaccination strategies targeting persons with *Mtb* infection to assess the absolute and relative impacts on annual TB incidence in the U.S.

## Methods

### Model Structure

#### Population strata and parameterization

The model stratifies the U.S. population into four mutually exclusive populations: persons living with HIV, persons with non-HIV medical comorbidities, non-U.S.-born persons, and U.S.-born persons. Because these groups overlap (e.g., a non-U.S.-born person living with HIV belongs to multiple risk categories), each individual is assigned to the population with the highest TB progression risk, in the following order: (1) persons living with HIV (PLWH), (2) persons with non-HIV medical comorbidities that increase TB progression risk, (3) non-U.S.-born persons without HIV or a non-HIV medical comorbidity that increases TB progression risk, and (4) U.S.-born persons without HIV or a non-HIV medical comorbidity that increases TB progression risk. Under this hierarchy, a non-U.S.-born person living with HIV would be assigned to the PLWH population because HIV confers the highest progression risk (Table S1).

#### Compartmental structure

We model *Mtb* infection and progression to TB using five main compartments. When a susceptible person acquires *Mtb* infection, they enter either a fast latent state (*L*_*f*_, representing individuals within approximately two years of initial infection who face the highest risk of rapid progression to TB disease) or a slow latent state (*L*_*s*_, representing individuals with established, stable infection who face a low annual risk of reactivation). Individuals in either latent state may progress to TB disease (infectious, *I*) and then be treated and recover (*R*) (Figure 1). New *Mtb* infections enter either *L*_*f*_ or *L*_*s*_ in proportions that are specific to each strata.^16,17^ Previously infected and recovered individuals receive partial protection against reinfection, so approximately one in five reinfection events leads to a new latent infection.^18^ Approximately 1 million people enter the non-U.S.-born population each year, and those with *Mtb* infection enter the slow latent state.^19^ Population size is held constant within each population. Full model equations are included in Appendix: Model Equations and Technical Details.

**Figure 1.**
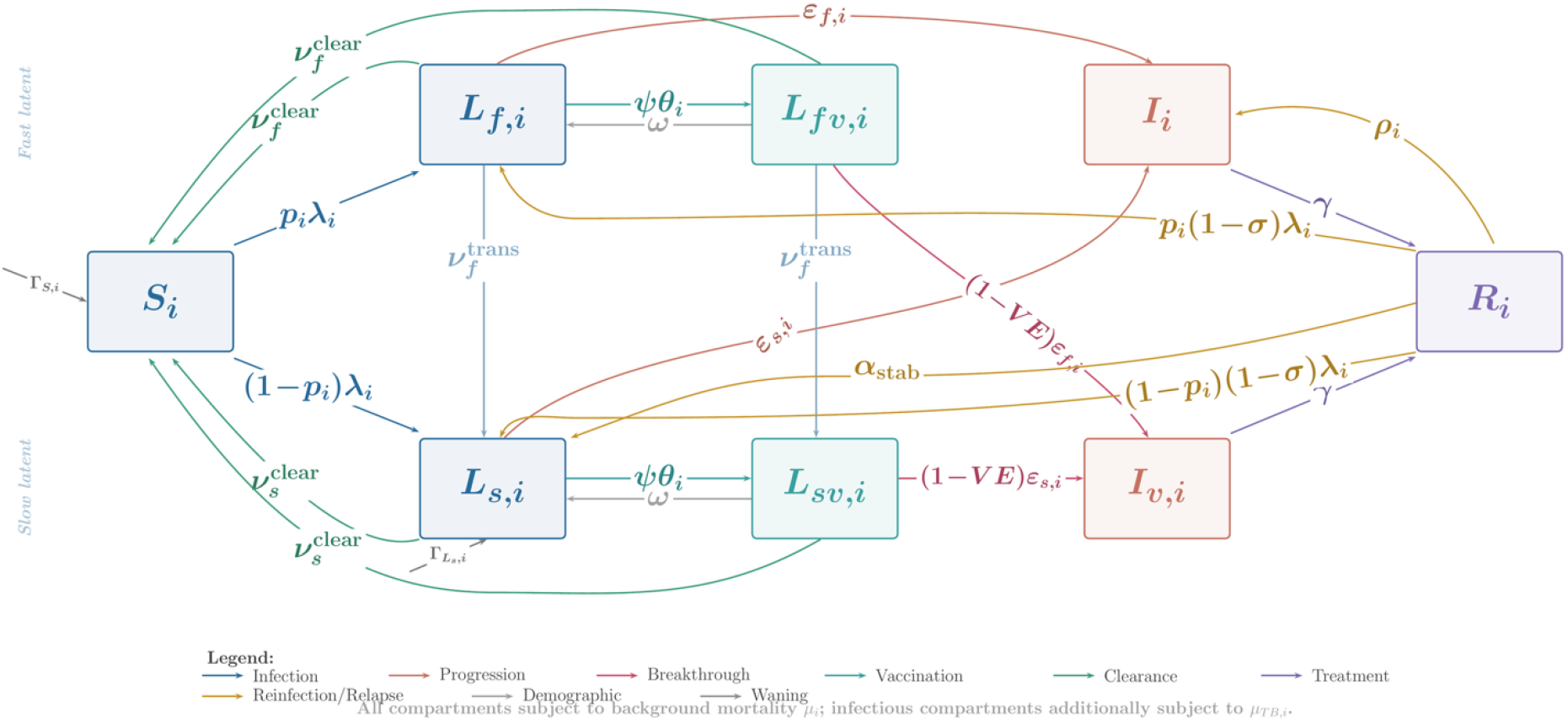
Compartmental model structure for stratum *i*. Each stratum contains eight compartments tracking susceptible (*S*), latent fast and latent slow infection (unvaccinated: *L*_*f*_, *L*_*s*_, vaccinated: *L*_*fv*_, *L*_*sv*_), infectious disease (unvaccinated: *I*, vaccinated: *I*_*v*_), and recovered (*R*) states. The structure is replicated across four risk strata for 32 ODEs total.

Slow reactivation rates (*ε*_*s*_) were based on published empirical estimates of reactivation risk by population group.^4^ Fast progression rates (*ε*_*f*_) were derived from prior modeling estimates of early progression risk following *Mtb* infection, which synthesize reported incidence of TB disease by time since infection from contact-tracing studies of persons with recent exposure^16^, with rates adjusted upward for PLWH and persons with non-HIV medical comorbidities to reflect faster progression to TB disease in these populations (full derivation in Appendix: Model Equations and Technical Details). All parameter values, ranges, and sources are provided in Table S2.

#### Force of infection and mixing

The force of infection (*λ*_*i*_) for each stratum depends on the prevalence of TB disease across all strata, weighted by a mixing matrix that controls the extent to which transmission occurs within the same population group versus between different groups (Appendix: Model Equations and Technical Details). For PLWH and persons with non-HIV medical comorbidities, we assumed that contact patterns with respect to *Mtb* transmission are proportional to population size (i.e., these groups do not have preferential within-group contact), since elevated TB risk in these groups likely arises from faster disease progression rather than differential exposure patterns. For U.S.-born and non-U.S.-born persons, we assumed that 70% of transmission-relevant contacts occur within the same nativity group (*ε*^*mix*^_*i*_*= 0·70*), based on genotyping evidence that TB transmission in the U.S. occurs predominantly within nativity groups.^2,20^ We varied this parameter in sensitivity analyses.

### Model Calibration

We calibrated the transmission rate (*β*, shared across strata) and four stratum-specific *Mtb* infection prevalences to 2024 TB incidence data (*n=10,388* reported cases).^1^ We minimized the sum of squared relative errors between model-predicted and observed equilibrium incidence. The full objective function and solver details are provided in Appendix: Model Equations and Technical Details.

The model was solved at equilibrium, consistent with near-stable U.S. TB incidence before the COVID-19 pandemic (1% per year decline, 2012–2019) and confirmed by forward integration (Sensitivity and Uncertainty Analyses). An alternative calibration that fixes *Mtb* infection prevalence and estimates reactivation rates instead is presented in Appendix: Alternative Calibration Analysis. We assessed model adequacy by comparing model outputs that were not used as calibration targets (such as the proportion of cases from recent transmission and reactivation) against independent surveillance data (Appendix: Model Calibration and Validation).

### Vaccination

Individuals with *Mtb* infection in targeted populations are vaccinated and move to vaccinated compartments (*L*_*fv*_ or *L*_*sv*_) where progression risk is reduced by the vaccine efficacy. Vaccine protection wanes over time, returning individuals to unvaccinated compartments. We modeled a prevention-of-disease vaccine that reduces progression risk but does not clear existing infection or prevent new *Mtb* infection. Vaccinated persons progressed to TB disease at a reduced rate compared with unvaccinated persons in the same risk group, with the size of the reduction set by vaccine efficacy (e.g., vaccinated persons progressed at half the rate of unvaccinated persons at 50% vaccine efficacy). Because the eventual characteristics of a licensed vaccine remain uncertain, we varied vaccine efficacy from 10–95% and duration of vaccine protection from 5–30 years in sensitivity and threshold analyses (Sensitivity and Uncertainty Analyses). Vaccine efficacies below approximately 30% are shown for completeness of the parameter space but fall below the range that would plausibly support licensure.

### Intervention Scenarios

We tested multiple vaccination strategies that varied in the populations targeted for vaccination (Table 1). Strategies ranged from single-group targeting (only PLWH or only those with non-HIV medical comorbidities) to multiple groups targeted (PLWH + Medical, PLWH + Non-U.S.-Born, All-High-Risk) to vaccinating all persons with *Mtb* infection in the U.S. (All *Mtb*-Infected). We evaluated each strategy under two sets of vaccine and coverage assumptions, referred to as the optimistic and plausible scenarios. The optimistic scenario used a vaccine efficacy (VE) estimate of 70% and a vaccination rate of 50% per year for all strategies, representing what could be achieved if a vaccine with higher efficacy becomes available and vaccination programs reach a large fraction of the target population each year. The plausible scenario used a VE estimate of 50%, 10-year duration of protection, and strategy-specific vaccination rates applied to *Mtb*-infected individuals within each targeted population: 5% per year for most strategies, 10% for PLWH (reflecting routine HIV care engagement),^21^ and 2% for persons with *Mtb* infection (reflecting the large eligible population and uncertainties about how to consistently reach this group). Both scenarios are evaluated at equilibrium for each strategy. All strategies assume, and do not explicitly model, prior testing for *Mtb* infection.

**Table 1.** Vaccination strategy definitions. Vaccination rates shown for both optimistic scenario (VE = 70%, 50%/yr for all strategies) and plausible scenario (VE = 50%, strategy-specific rates). 10-year duration of protection in both scenarios. PLWH = persons living with HIV. ESRD = end-stage renal disease. Shorthand labels used in subsequent tables and figures are shown in parentheses.

| Strategy (shorthand) | Vaccination rate (optimistic) | Vaccination rate (plausible) | Target population |
| --- | --- | --- | --- |
| PLWH, non-U.S.-born persons, and persons with medical comorbidities<br>(**All-High-Risk**) | 50%/yr | 5%/yr | PLWH, persons with medical comorbidities that increase TB progression risk (i.e., diabetes, ESRD, immunosuppressive therapy, and solid organ transplant), and all non-U.S.-born persons |
| PLWH and non-U.S.-born persons<br>(**PLWH + Non-U.S.-Born**) | 50%/yr | 5%/yr | PLWH and non-U.S.-born persons, including non-U.S.-born persons with non-HIV medical comorbidities. Does not include U.S.-born persons with medical comorbidities |
| All *Mtb*-infected persons<br>(**All *Mtb*-Infected**) | 50%/yr | 2%/yr | All *Mtb*-infected persons across all four populations, including U.S.-born persons |
| PLWH and persons with medical comorbidities<br>(**PLWH + Medical**) | 50%/yr | 5%/yr | PLWH and persons with medical comorbidities that increase TB progression risk |
| Persons with non-HIV medical comorbidities<br>(**Medical comorbidities**) | 50%/yr | 5%/yr | Persons with medical comorbidities that increase TB progression risk, including diabetes, ESRD, immunosuppressive therapy, and solid organ transplant |
| **PLWH** | 50%/yr | 10%/yr | Persons living with HIV |

### Outcome Measures

We calculated three primary outcomes at equilibrium (approximately 50 years of forward integration): annual cases prevented, percent reduction in TB incidence relative to the baseline, no-vaccine scenario, and number needed to vaccinate (NNV = annual vaccinations / annual TB cases prevented), an inverse measure of vaccination efficiency. All outcome measures were computed per population and overall. We decomposed total vaccine impact into direct effects (TB cases prevented in vaccinated individuals) and indirect effects (TB cases prevented in unvaccinated individuals through reduced transmission). We estimated direct effects by running the model with the force of infection held fixed at baseline levels, and total effects by allowing the force of infection to change as vaccination reduced TB prevalence. The difference between these runs gave the indirect effect contribution (Appendix: Stratum-Level Impact and Direct and Indirect Vaccine Effects). We calculated 95% uncertainty intervals as the 2·5th and 97·5th percentiles across 500 Latin Hypercube Sampling draws that simultaneously varied 12 parameters (described below).

### Sensitivity and Uncertainty Analyses

To quantify uncertainty in model estimates, we performed 500 Latin Hypercube Sampling (LHS) draws under each scenario across 12 parameters spanning TB natural history and vaccine characteristics (Table S3), varying all parameters simultaneously with full model re-calibration at each draw, and derived 95% uncertainty intervals and Partial Rank Correlation Coefficients (PRCC).^22^ To assess the influence of individual parameters, we separately varied 11 natural history parameters one at a time with re-calibration at each value (Appendix: Sensitivity and Uncertainty Analysis). We varied the annual vaccination rate from 1 to 50% per year for each strategy and reported effective coverage, defined as the percentage of *Mtb*-infected individuals vaccinated at equilibrium (Appendix: Vaccination Coverage-Impact Analysis). Because the primary analysis reports long-run impact at equilibrium, we also integrated the model forward over 30 years to assess how quickly vaccination impact accumulates after program initiation.

All analyses were implemented in R (v4·5·2) using deSolve for ordinary differential equations (ODE) integration, optim for model calibration, and lhs for Latin Hypercube Sampling.

## Results

### Model Calibration and Validation

The model was calibrated to 2024 TB incidence data^1^ (n=10,388 reported cases). The model reproduced the proportions of TB cases from recent transmission (11·1% modeled vs. 10–15% from NTSS genotyping data), reactivation (86·4% modeled vs. 85–90% from NTSS), and post-treatment relapse (2·5% modeled vs. 3–6% from NTSS) without these proportions being used as calibration targets (Figure S1).^2,23^

### Strategy Comparison

Targeting PLWH, non-U.S.-born persons, and persons with non-HIV medical comorbidities (All-High-Risk) prevented the most cases under both scenarios (Table 2, Figure 2). Under the optimistic scenario, vaccinating all high-risk groups prevented 5,385 cases per year (95% UI: 3,589–6,939 cases prevented, 51·8% reduction, NNV = 366, 95% UI: 205–862) and vaccinating all those with *Mtb* infection (All *Mtb*-infected) prevented 6,008 cases (57·8% reduction, NNV = 363) (Table 2). While absolute impact was different, the rank order for each strategy in terms of programmatic efficiency (NNV) and relative impact (proportion annual cases prevented) were the same under both the plausible and optimistic scenarios. NNV estimates were lower under the optimistic scenario because higher VE prevents more cases per vaccinated person. Effective coverage among targeted populations reached 79–81% under the optimistic scenario, compared with 13–46% under the plausible scenario.

**Table 2.** Vaccination strategy comparison under optimistic (VE = 70%, vaccination rate = 50%/yr) and plausible (VE = 50%, strategy-specific annual vaccination rate) scenarios with 10-year protection. Optimistic scenario: VE = 70%, 10-year duration of protection, vaccination rate = 50%/yr for all strategies. Plausible scenario: VE = 50%, 10-year duration of protection, strategy-specific vaccination rates (5%/yr for most strategies, 10%/yr for PLWH, 2%/yr for All *Mtb*-Infected). Strategies ordered by cases prevented under the optimistic scenario. Cases Prevented and NNV shown as point estimates from the calibrated deterministic model. 95% uncertainty intervals (UI) are the 2·5th and 97·5th percentiles across 500 Latin Hypercube Sampling draws that simultaneously varied 12 model parameters. Point estimates fall within UIs for all strategies and both scenarios. NNV = number needed to vaccinate. Effective coverage is the percentage of *Mtb*-infected individuals within targeted populations that are in vaccinated compartments at equilibrium.

| Strategy | Optimistic Scenario |  |  |  | Plausible Scenario |  |  |  |
| --- | --- | --- | --- | --- | --- | --- | --- | --- |
|  | Cases Prevented (95% UI) | % Reduction | NNV (95% UI) | Effective Coverage | Cases Prevented (95% UI) | % Reduction | NNV (95% UI) | Effective Coverage |
| All *Mtb*-Infected | 6,008<br>(4,143–7,653) | 57.8% | 363 (204–832) | 79.5% | 739 (199–1,133) | 7.1% | 507 (283–1,998) | 13.2% |
| All-High-Risk | 5,385<br>(3,589–6,939) | 51.8% | 366 (205–862) | 79.3% | 1,348<br>(527–2,011) | 13.0% | 510 (284–1,631) | 27.5% |
| PLWH + Non-U.S.-Born | 4,331<br>(2,746–5,674) | 41.7% | 377 (216–861) | 65.2% | 925 (299–1,395) | 8.9% | 537 (313–1,875) | 19.6% |
| PLWH + Medical | 2,473<br>(1,598–3,174) | 23.8% | 291 (148–919) | 79.8% | 617 (152–946) | 5.9% | 411 (203–2,301) | 28.2% |
| Medical comorbidities | 2,223<br>(1,419–2,847) | 21.4% | 319 (164–1,022) | 79.8% | 552 (116–851) | 5.3% | 451 (221–2,653) | 28.1% |
| PLWH | 264 (19–394) | 2.5% | 46 (18–806) | 81.3% | 99 (4–239) | 1.0% | 68 (19–1,674) | 46.1% |

**Figure 2.**
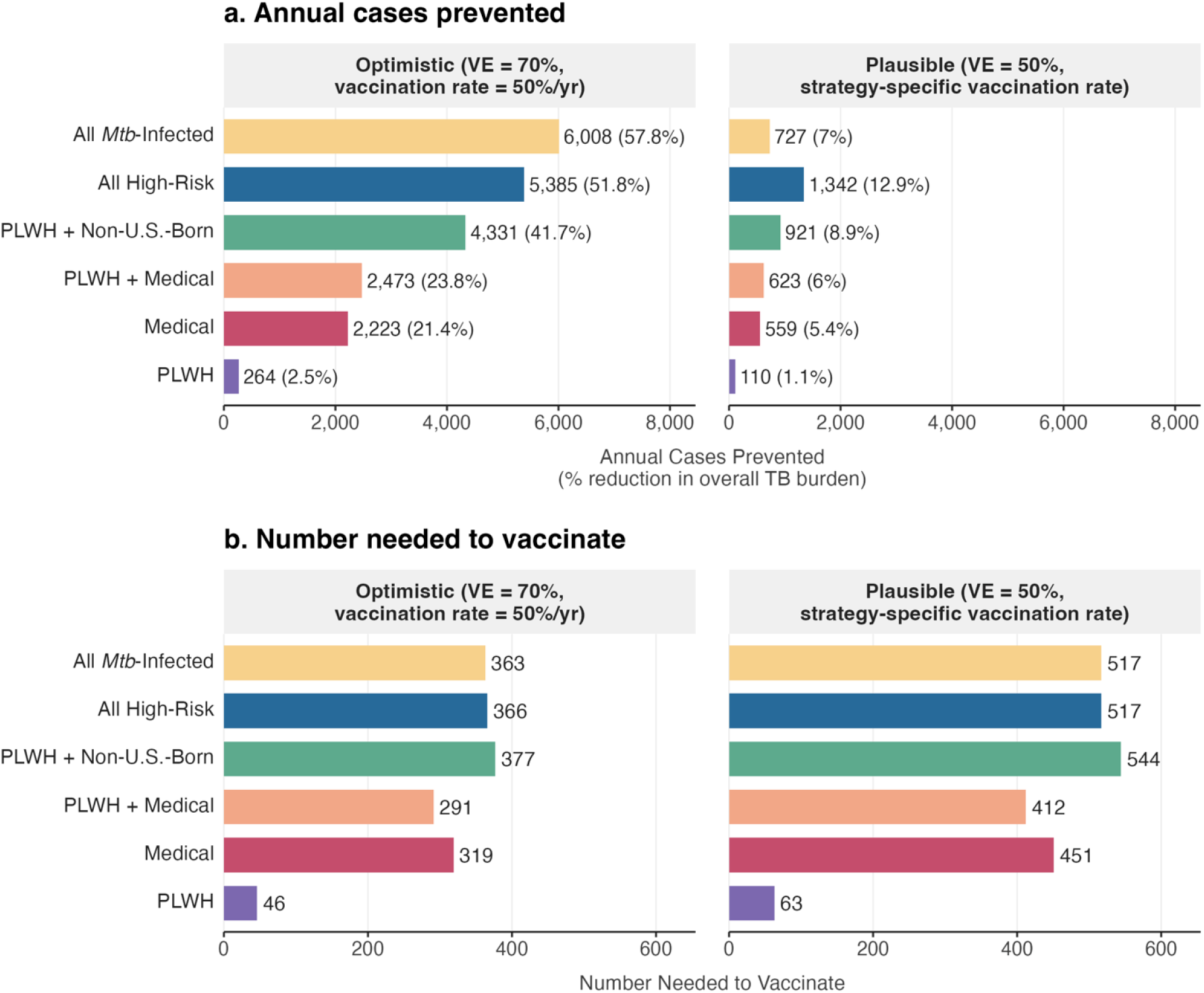
Vaccination strategy comparison across scenarios. Panel a shows annual cases prevented (with percent reduction in overall TB burden in parentheses). Panel b shows the number needed to vaccinate. Within each panel, the left facet shows results under the optimistic scenario (VE = 70%, 10-year duration of vaccine protection, vaccination rate = 50%/yr for all strategies) and the right facet shows results under the plausible scenario (VE = 50%, 10-year duration of vaccine protection, strategy-specific vaccination rates: 5%/yr for most strategies, 10%/yr for PLWH, 2%/yr for All *Mtb*-Infected). Strategies are ordered by cases prevented under the optimistic scenario. Population-level results by stratum are provided in Figure S2. Corresponding numerical values with 95% uncertainty intervals from Latin Hypercube Sampling are provided in Table 2.

Under the plausible scenario, vaccinating all high-risk groups prevented 1,348 cases per year (95% UI: 527–2,011 cases prevented, 13·0% reduction, NNV = 510, 95% UI: 284–1,631), while targeting only PLWH had the lowest NNV (68), indicating the highest programmatic efficiency of any scenario, but prevented only 100 cases annually (1·0% reduction). Strategies that included non-U.S.-born persons prevented 926 to 1,348 cases per year (8·9–13·0% reduction), compared with 100 to 617 cases (1·0–5·9%) for strategies that targeted population subgroups defined by clinical characteristics (PLWH, non-HIV medical comorbidities).

### Direct and Indirect Effects

Across strategies, direct effects accounted for 77–89% of total impact under both scenarios (Table S4). Indirect effects were proportionally largest for PLWH (23% of impact). When vaccinating all groups at high risk, targeted risk groups showed approximately uniform percent reductions in cases across scenarios: under the optimistic scenario, 56·7% for PLWH, 56·6% for persons with non-HIV medical comorbidities, and 56·8% for non-U.S.-born persons, under the plausible scenario, 14·8% for PLWH, 14·3% for persons with non-HIV medical comorbidities, and 14·1% for non-U.S.-born persons (Figure S2). These groups differ by more than 30-fold in population size and nearly 4-fold in TB incidence rate (Table S1). The most absolute TB cases prevented were among non-U.S.-born persons (2,691 of 5,385 under optimistic, 669 of 1,348 under plausible) (Figure S2). Despite receiving no vaccination, U.S.-born persons received a 22·4% reduction in cases (323 cases) under the optimistic scenario and a 5·5% reduction (79 cases) under the plausible scenario through indirect vaccine effects on transmission (Figure S2).

### Vaccine Characteristic Requirements

Because the characteristics of a licensed vaccine remain uncertain, we identified the minimum vaccine efficacy and duration of protection required to achieve policy-relevant reductions in cases. Under the optimistic scenario, vaccinating all high-risk groups (All-High-Risk) would achieve a ≥20% reduction in annual TB cases with a vaccine of 30% efficacy and 10-year duration of protection, a ≥35% reduction at 50% efficacy, and would halve annual cases at 70% efficacy (Figure 3). Under the plausible scenario, the same strategy would achieve ≥5%, ≥10%, and ≥15% reductions at vaccine efficacies of 20%, 40%, and 60% respectively at 10-year duration of protection. Other strategies required higher efficacy to reach the same thresholds under both scenarios. Extending the duration of vaccine protection from 10 to 20 years reduced the minimum efficacy required by 5–20 percentage points across strategies and thresholds (Figure 3). Vaccinating only PLWH could not reach the tested thresholds at any efficacy or duration under either scenario because the PLWH population is too small, despite its low NNV.

**Figure 3.**
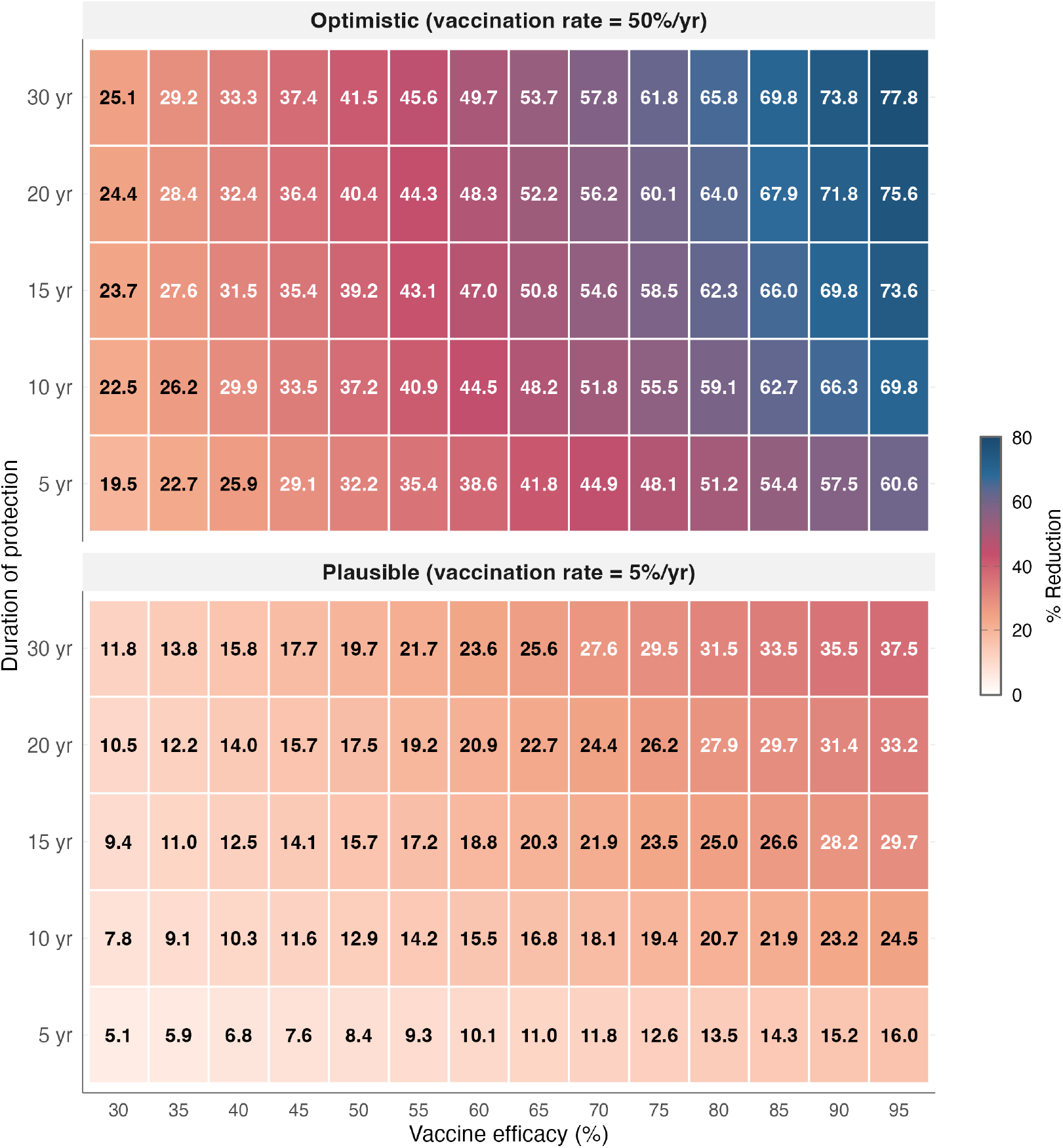
Vaccine efficacy vs. duration of protection: target product characteristics. Percent reduction in annual TB cases for the All-High-Risk strategy across VE (30–95%) and duration of vaccine protection (5–30 years). These results characterize the vaccine properties needed to achieve specific levels of population impact under the optimistic scenario (50%/yr vaccination rate, left panel) and the plausible scenario (5%/yr vaccination rate, right panel). At the M72/AS01_E_ reference point (VE = 50%, 10 yr), reduction = 12·9%.

## Discussion

We evaluated multiple vaccination strategies for adults with *Mtb* infection in the U.S. using a mathematical model of TB transmission calibrated to 2024 incidence. Under an optimistic scenario combining a vaccine of 70% efficacy with an annual vaccination rate of 50%, vaccinating all high-risk groups reduced annual TB cases by more than half (52%, NNV = 366). Under a more conservative scenario of 50% efficacy and a 5% annual vaccination rate, the same strategy reduced annual cases by 13%. Across scenarios, no single strategy was best on both absolute impact and efficiency. Vaccinating all high-risk groups prevented the most cases, while vaccinating only PLWH was the most efficient per dose but prevented fewer cases overall. Any strategy that produced substantial population-level impact included non-U.S.-born persons.

To maximize impact and efficiency, a TB vaccination strategy in the U.S. should include non-U.S.-born persons and PLWH. Non-U.S.-born persons account for approximately 77% of U.S. TB cases, so any strategy that excludes this group forgoes the majority of achievable vaccine impact. Vaccinating PLWH and non-U.S.-born persons (PLWH + Non-U.S.-Born) captured 69% of the impact of vaccinating all high-risk groups with 72% of the vaccinations, preventing 4,331 cases per year (41·7% reduction, NNV = 377) under the optimistic scenario and 926 cases (8·9% reduction, NNV = 537) under the plausible scenario. Among clinical risk groups, targeting PLWH alone was substantially more efficient per dose than targeting persons with non-HIV medical comorbidities (NNV = 68 vs 451 under the plausible scenario). PLWH is a small population with sharply elevated TB progression risk, while the much larger medical-comorbidities population has only modestly elevated risk, so each dose delivered to the broader group prevents fewer cases.

Existing clinical services can reach PLWH through routine HIV care, and immigration-related medical examinations and post-arrival health screening can reach non-U.S.-born persons. Reaching non-U.S.-born persons outside of clinical settings would require expanding *Mtb* infection screening through community-based program. All strategies we modeled assume *Mtb* infection screening is already in place, so achievable impact depends on screening uptake. Achievable vaccination rates will depend on the capacity of each delivery platform to identify and vaccinate eligible individuals. Combining TB vaccination with expanded *Mtb* infection testing and treatment could increase impact beyond what either intervention achieves alone.

Because most U.S. TB cases arise from reactivation of existing *Mtb* infection rather than recent transmission, vaccine impact depends mainly on how many infected individuals the program vaccinates. Indirect effects on transmission to unvaccinated populations were modest but not negligible and were largest in proportional terms when vaccinating PLWH. Halving annual TB cases by vaccinating all high-risk groups would require a vaccine of at least 70% efficacy at 10-year duration of protection, within the range of current M72/AS01_E_ Phase IIb estimates, combined with a 50% annual vaccination rate in targeted populations. Longer durations of protection relaxed this requirement, reducing the minimum required efficacy to 65% at 20 or more years of protection. The M72/AS01_E_ Phase III trial will establish efficacy in a high-burden setting that may not generalize to the U.S., and its approximately three-year follow-up will leave duration of protection unknown at licensure. Our results show that targeted vaccination can deliver meaningful reductions in U.S. TB incidence across the plausible range of duration assumptions.

Our NNV estimates fall within the range reported for other recommended adult vaccines in the United States and for hypothetical TB vaccine programs modeled in other settings (Figure S5). For comparison, approximately 777 adults aged ≥65 years need to be vaccinated to prevent one influenza-related hospitalization,^24^ and approximately 3,333 need to be vaccinated to prevent one hospitalization from invasive pneumococcal disease.^25^ Prior modeling of older-adult (aged 60–64) TB vaccination in China reported NNV estimates ranging from 230 to 1,022 at 60% vaccine efficacy and 10-year duration of protection over a 25-year time horizon.^26^

This analysis has several limitations. The equilibrium assumption is a simplification. If U.S. TB incidence resumes its pre-pandemic decline, the model would overestimate long-term impact. The model does not include age structure, which limits analysis of age-targeted strategies. Current TB vaccine trials enroll adults aged 15–45, and pediatric TB accounts for a small proportion of U.S. cases. We assume uniform vaccine efficacy across populations, although immunocompromised individuals may have reduced vaccine response, lowering the population-level impact of clinical-only strategies. To our knowledge, these are the first estimates comparing TB vaccination strategies across clinically and demographically defined high-risk populations in a low-burden setting. Future research should evaluate the cost-effectiveness of these strategies under different implementation scenarios, including comparisons with *Mtb* infection testing and preventive therapy, which may accompany vaccination as part of ongoing U.S. TB elimination efforts.

## Data Availability

All model code, calibration routines, sensitivity analyses, and figure-generating scripts are available at https://github.com/jrothman27/tb-vaccination-model.

## Acknowledgements

This study was funded by an American Lung Association Catalyst Award (CA-943920), an NIH K01 (NIH/NIAID K01AI166093-01A1), and a developmental grant from the NIH Center for AIDS Research at Emory University (P30 AI050409) to KNN. This work was also supported by an NIH K24 (K24 AI114444) to NRG and the Emory/Georgia Tuberculosis Research Advancement Center (P30 AI168386). The lead author (JER) was supported by the Ferguson Fellowship. The funders of this study had no role in its design and conduct, nor the analysis and presentation of results.

