## Supplemental Materials for "Targeted Tuberculosis (TB) Vaccination Strategies in the United States: A Modeling Study"

**Model Validation** **5**

**Reference Tables** **7**

**Direct and Indirect Vaccine Effects** **9**

**Sensitivity and Uncertainty Analysis** **10**

One-Way Sensitivity . . . . . . . . 10
Latin Hypercube Sampling and PRCC . . . . . . . . 11

**Vaccination Coverage-Impact Analysis** **11**

**Time-to-Impact Analysis** **14**

**NNV Benchmarking** **16**

### Model Equations and Technical Details

#### Complete ODE System

The model consists of eight ordinary differential equations per stratum , for 32 equations total. For stratum :

$$
\begin{aligned}
\frac{dS_i}{dt} &= \Gamma_{S,i} - \lambda_i S_i - \mu_i S_i + \nu_f^{\text{clear}} L_{f,i} + \nu_s^{\text{clear}} L_{s,i} + \nu_f^{\text{clear}} L_{fv,i} + \nu_s^{\text{clear}} L_{sv,i} \\
\frac{dL_{f,i}}{dt} &= p_i \lambda_i S_i + p_i \lambda_i R_i (1-\sigma) - \varepsilon_{f,i} L_{f,i} - \nu_f^{\text{trans}} L_{f,i} - \nu_f^{\text{clear}} L_{f,i} \\
&\quad - \psi \theta_i L_{f,i} + \omega L_{fv,i} - \mu_i L_{f,i} \\
\frac{dL_{s,i}}{dt} &= \Gamma_{L_s,i} + \nu_f^{\text{trans}} L_{f,i} + (1-p_i)\lambda_i S_i + (1-p_i)\lambda_i R_i (1-\sigma) + \alpha_{\text{stab}} R_i \\
&\quad - \varepsilon_{s,i} L_{s,i} - \nu_s^{\text{clear}} L_{s,i} - \psi \theta_i L_{s,i} + \omega L_{sv,i} - \mu_i L_{s,i} \\
\frac{dL_{fv,i}}{dt} &= \psi \theta_i L_{f,i} - (1-\text{VE})\varepsilon_{f,i} L_{fv,i} - \nu_f^{\text{trans}} L_{fv,i} - \nu_f^{\text{clear}} L_{fv,i} - \omega L_{fv,i} - \mu_i L_{fv,i} \\
\frac{dL_{sv,i}}{dt} &= \psi \theta_i L_{s,i} + \nu_f^{\text{trans}} L_{fv,i} - (1-\text{VE})\varepsilon_{s,i} L_{sv,i} - \nu_s^{\text{clear}} L_{sv,i} - \omega L_{sv,i} - \mu_i L_{sv,i} \\
\frac{dI_i}{dt} &= \varepsilon_{f,i} L_{f,i} + \varepsilon_{s,i} L_{s,i} + \rho_i R_i - \gamma I_i - (\mu_i + \mu_{\text{TB},i}) I_i \\
\frac{dI_{v,i}}{dt} &= (1-\text{VE})\varepsilon_{f,i} L_{fv,i} + (1-\text{VE})\varepsilon_{s,i} L_{sv,i} - \gamma I_{v,i} - (\mu_i + \mu_{\text{TB},i}) I_{v,i} \\
\frac{dR_i}{dt} &= \gamma I_i + \gamma I_{v,i} - \lambda_i R_i (1-\sigma) - \rho_i R_i - \alpha_{\text{stab}} R_i - \mu_i R_i
\end{aligned}
$$

#### Mixing Matrix

The mixing matrix element governing the force of infection (Force of infection and mixing) is defined as:

$$
M_{ij} = \begin{cases} \varepsilon_i^{\text{mix}} + (1-\varepsilon_i^{\text{mix}}) \cdot \dfrac{N_j}{\sum_k N_k} & \text{if } i = j \\ (1-\varepsilon_i^{\text{mix}}) \cdot \dfrac{N_j}{\sum_k N_k} & \text{if } i \neq j \end{cases}
$$

where is the assortative mixing parameter for stratum . We set (proportionate mixing) for the PLWH and medical comorbidities strata and (assortative mixing) for the non-U.S.-born and U.S.-born strata. The parameter is applied symmetrically to both nativity strata because if non-U.S.-born persons have disproportionate contact within their communities, then U.S.-born persons also have disproportionately more contact with other U.S.-born persons.

#### Demographic Balance

The recruitment rates that maintain constant population size within each stratum are:

$$\Gamma_{S,i} = (\mu_i + \iota_i) N_i \cdot (1 - \text{mtb\_prev}_i)$$

$$\Gamma_{L_s,i} = (\mu_i + \iota_i) N_i \cdot \text{mtb\_prev}_i$$

where $\mu_i$ is the background mortality rate, $\iota_i$ is the immigration rate (nonzero only for the non-U.S.-born stratum, *ι=0·023*/yr),[1] and $\text{mtb\_prev}_i$ is the *Mtb* infection prevalence among new entrants (a calibrated parameter).

**Fast Progression Rate Derivation**

Individuals exit the fast latent compartment ($L_f$) through three competing pathways: progression to TB disease ($\varepsilon_f$), stabilization to slow latent infection ($\nu_f^{trans}$), and background mortality ($\mu$). The $L_f$ clearance rate ($\nu_f^{clear}=1{\cdot}4\times10^{-6}$/yr) is negligible and omitted. The mean residence time is:

$$T_{L_f} = \frac{1}{\varepsilon_f + \nu_f^{\text{trans}} + \mu}$$

Setting years[2] and solving for :

$$\varepsilon_{f,\text{base}} = \frac{1}{T_{L_f}} - \nu_f^{\text{trans}} - \mu = \frac{1}{2} - 0{\cdot}30 - 0{\cdot}01 = 0{\cdot}19 \text{ /year}$$

Stratum-specific values were obtained by applying fixed relative risk multipliers:

$$\varepsilon_{f,i} = \varepsilon_{f,\text{base}} \times \text{RR}_i$$

**Fast progression rate multipliers by stratum.**

| Stratum | *RR* | $\varepsilon_f$ (/yr) | Implied $T_{Lf}$ (yr) | Rationale |
|---|---|---|---|---|
| PLWH | 2·50 | 0·475 | 1·27 | Immunosuppression accelerates progression |
| Medical comorbidities | 1·25 | 0·238 | 1·86 | Moderate risk elevation (diabetes, ESRD) |
| Non-U.S.-born | 1·00 | 0·190 | 2·00 | Baseline immunocompetent |
| U.S.-born | 1·00 | 0·190 | 2·00 | Baseline immunocompetent |

$T_{Lf}$ computed as $1/(\varepsilon_f+\nu_f^{trans}+\mu_i)$ with stratum-specific $\mu_i$. The RR multipliers are modeling assumptions. Their influence on model outputs is assessed in sensitivity analysis (Sensitivity and Uncertainty Analysis). The proportion entering fast latency ($p$) was also adjusted upward for HIV ($p=0.25$) and Medical ($p=0.20$) relative to NUSB/USB ($p=0.15$).[3,4]

The model's emergent fast-progression fraction (11·1% of cases from →I) was not a calibration target but serves as a validation metric, consistent with NTSS genotyping estimates of 10–15% recent transmission.[5,6]

**Mechanism of Vaccine Action**

Vaccination moves individuals from unvaccinated latent compartments (, ) to vaccinated counterparts (, ) at rate , where is the annual vaccination rate and for targeted strata and 0 otherwise. In vaccinated compartments, the progression rate to TB disease is reduced by a factor of . This multiplicative reduction applies equally to fast and slow latent compartments. Vaccine protection wanes at rate , returning individuals from vaccinated to unvaccinated compartments (, ). New infections acquired after vaccination enter the standard unvaccinated or compartments, not the vaccinated compartments.

Vaccine protection wanes at rate $\omega=1/duration$, returning individuals from vaccinated to unvaccinated compartments ($L_{fv}\rightarrow L_f$, $L_{sv}\rightarrow L_s$). Susceptible ($S$) and recovered ($R$) individuals are not vaccinated because the vaccine acts on existing *Mtb* infection. New infections acquired after vaccination enter the standard unvaccinated $L_f$ or $L_s$ compartments, not the vaccinated compartments.

**Calibration Objective Function**

The transmission rate and stratum-specific *Mtb* infection prevalences among recruits (mtb_prev) were calibrated by minimizing the sum of squared relative errors between model-predicted and observed equilibrium incidence:

$$\text{SSE} = \sum_{i=1}^{4} \left( \frac{\text{Inc}_i^{\text{model}} - \text{Inc}_i^{\text{target}}}{\text{Inc}_i^{\text{target}}} \right)^2$$

The optimization used a quasi-Newton optimization algorithm with bounded constraints (L-BFGS-B) and log-transformed parameters (optim() in R).[7] To estimate equilibrium, the ODE system was integrated for 50 years using the LSODA solver (deSolve package), reaching convergence at and population conservation verified to .

The calibrated parameter values and incidence targets are shown in the calibration summary table below. The model reproduced all four stratum-specific incidence targets with <0·5% error, which is expected because five free parameters ($\beta$ and four mtb_prev) are fit to four targets. Model adequacy is instead assessed through validation against independent data not used in calibration (Model Validation).

**Calibrated parameter values.** Five free parameters ($\beta$ and four stratum-specific *Mtb* infection prevalences) were calibrated to four incidence targets from 2024 NTSS data.

| Parameter | Calibrated Value | Description |
|---|---|---|
| $\beta$ | 5·70/yr | Transmission rate (shared across strata) |
| mtb_prev$_{PLWH}$ | 11·2% | *Mtb* infection prevalence among PLWH recruits |
| mtb_prev$_{Med}$ | 18·1% | *Mtb* infection prevalence among medical comorbidities recruits |
| mtb_prev$_{NUSB}$ | 30·2% | *Mtb* infection prevalence among non-U.S.-born recruits |
| mtb_prev$_{USB}$ | 0·86% | *Mtb* infection prevalence among U.S.-born recruits |
| PLWH | 34·83 per 100k | 417 cases/yr (*n* = 1·2M) |
| Medical comorbidities | 9·35 per 100k | 3,739 cases/yr (*n* = 40·0M) |
| Non-U.S.-born | 11·67 per 100k | 4,737 cases/yr (*n* = 40·6M) |
| U.S.-born | 0·57 per 100k | 1,444 cases/yr (*n* = 253·4M) |
| **Total** | **3·08 per 100k** | **10,337 cases/yr** |

All targets reproduced with *<0.1%* error. The mtb_prev parameters represent the fraction of new entrants to each stratum who have *Mtb* infection and are placed in $L_s$ upon entry. The NUSB value (30·2%) reflects immigration from higher-burden countries. The USB value (0·86%) reflects the low background transmission rate in the U.S.-born population.

#### Outcome Measure Definitions

Annual cases prevented, percent reduction, and NNV are defined as:

$$\text{Cases prevented} = \sum_i \left( I_i^{\text{baseline}} + I_{v,i}^{\text{baseline}} \right) - \sum_i \left( I_i^{\text{intervention}} + I_{v,i}^{\text{intervention}} \right)$$

where is the number of infectious individuals in stratum who were unvaccinated at the time of progression, is the number of infectious individuals in stratum who were vaccinated (i.e., progressed despite vaccination), and the summation is over all four strata ( PLWH, Medical comorbidities, Non-U.S.-born, U.S.-born). Superscripts indicate the scenario: baseline (no vaccination) or intervention (with vaccination). All quantities are evaluated at equilibrium.

*Percent reduction=Cases prevented/i(*$I_i^{baseline}+I_{v,i}^{baseline}$*)×100*

*NNV=Annual vaccinations administered/Annual cases prevented*

All quantities are evaluated at equilibrium. Because the full dynamic model is used for both baseline and intervention scenarios, these metrics capture both direct effects (reduced progression in vaccinated individuals) and indirect effects (reduced transmission to unvaccinated individuals).

### Alternative Calibration Analysis

#### Rationale

The primary calibration allows stratum-specific *Mtb* infection prevalences to float as free parameters, constrained only by the requirement that the model reproduce TB incidence targets. This yields a total equilibrium *Mtb* infection reservoir of 21·7 million ($L_f$+$L_s$+$L_{fv}$+$L_{sv}$, summed across strata), larger than the 12·6 million estimated from published reactivation rates and surveillance data. The difference arises because the compartmental model's slow latent compartment loses individuals continuously through immune clearance ($\nu_s^{clear}$), background mortality ($\mu_i$), and demographic turnover. The simpler cross-sectional formula does not account for these outflows:

$$\textit{Mtb}\text{ infection}_i = \frac{\text{Cases}_i \times p_{\text{reactivation},i}}{\varepsilon_{s,i}}$$

where cases and the proportion from reactivation are from NTSS surveillance data,[8] and $\varepsilon_{s,i}$ are the Ekramnia et al. (2024)[9] rates used in the primary calibration. The same number of TB cases can be produced by a larger latent reservoir with low reactivation rates (as in the primary calibration) or by a smaller reservoir with higher reactivation rates (as in the alternative calibration). These two approaches are not distinguishable from incidence data alone. The alternative calibration tests whether this choice affects vaccination impact estimates.

#### Method

In the alternative calibration, *Mtb* infection prevalence was held fixed at the estimates calculated from published reactivation rates and surveillance data, and stratum-specific reactivation rates ($\varepsilon_{s,i}$) were freed as calibrated parameters. The optimizer was constrained to match the same four TB incidence targets[8] as the primary calibration. All other parameters were held at primary calibration values.

#### Results

The alternative calibration produced reactivation rates 1·4–2·6× higher than the Ekramnia empirical rates,[9] compensating for the smaller *Mtb* infection reservoir. Despite this, intervention results were nearly identical: cases prevented differed by <2%, and strategy rankings were unchanged. The NNV for PLWH improved from 68 to 30 under the alternative calibration (a 2·3× reduction), because the smaller *Mtb* infection reservoir yields fewer annual vaccinations for the same stratum size. The primary calibration overestimates NNV more for some strata than others: HIV shows the largest NNV improvement (2·3×), Medical and Non-U.S.-born show moderate improvement (1·3–1·5×), and U.S.-born shows minimal change.

### Model Validation

We assessed model adequacy by comparing model outputs not used in calibration against independent data (Figure S1).

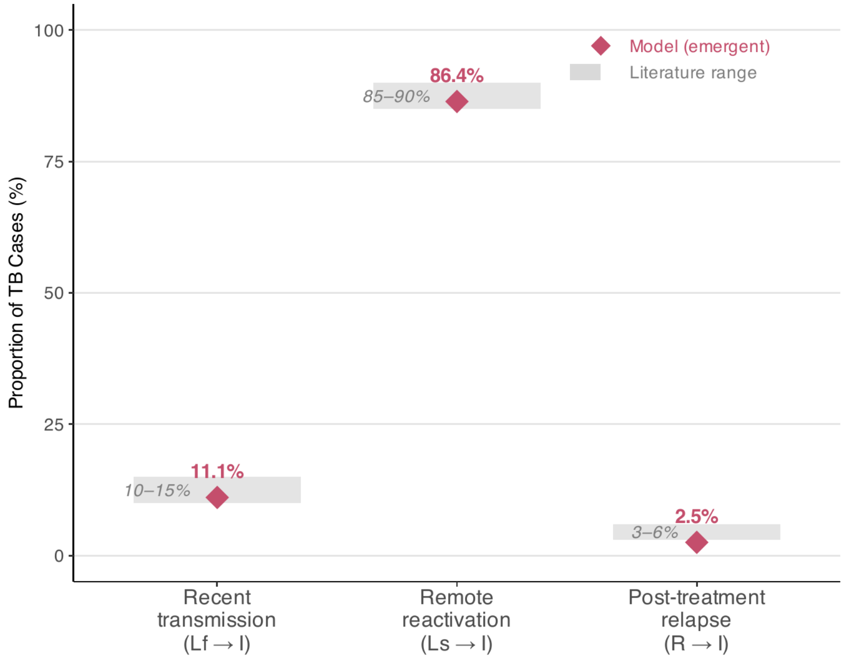

**Figure S1. Case source validation.** Model-produced proportions of TB cases from recent transmission ($L_f \rightarrow I$), remote reactivation ($L_s \rightarrow I$), and post-treatment relapse ($R \rightarrow I$) compared with published literature ranges. Literature ranges were 10–15% for recent transmission,[5,6] 85–90% for remote reactivation,[5,10] and 3–6% for post-treatment relapse.[11,12] These proportions were not used in calibration.

The model matched overall and non-U.S.-born recent transmission proportions well (11·1% vs. 13·0% overall, 8·2% vs. 7·8% for non-U.S.-born). Two stratum-level divergences are structurally explained. The U.S.-born stratum overestimates recent transmission because its small latent slow pool means Lf contributes disproportionately. The PLWH stratum underestimates recent transmission because the model defines PLWH by clinical status rather than transmission network. These divergences do not affect vaccination impact estimates because: (1) the U.S.-born stratum accounts for less than 14% of total cases and receives minimal direct vaccine impact, and (2) the PLWH stratum is small (417 cases per year), and its vaccination impact is driven by progression rates rather than transmission patterns. The model's *Mtb* infection reservoir (21·7 million) exceeds cross-sectional estimates (12·6 million) because the dynamic equilibrium requires a larger standing stock to offset competing outflows (clearance, mortality, demographic turnover) beyond reactivation alone. As shown in the alternative calibration analysis (Appendix: Alternative Calibration Analysis), fixing the reservoir at the smaller estimate and recalibrating reactivation rates produces nearly identical strategy rankings and cases prevented (within 2%), so this divergence does not affect the comparative conclusions.

### Reference Tables

**Table S1. Population strata.** PLWH = persons living with HIV. ESRD = end-stage renal disease. Incidence is TB disease incidence per 100,000 persons per year from baseline equilibrium of the calibrated model, which reproduces 2024 NTSS stratum-specific incidence targets within 0·1% error. [a]The Medical comorbidities stratum is dominated by diabetes (38·4M, 96% of stratum), with smaller contributions from ESRD, TNF-α inhibitor use, solid organ transplant, and other immunosuppressive conditions. The 40·0M figure reflects gross totals ( 41·3M) net of estimated overlap ( 890K). Diabetes prevalence is from the CDC National Diabetes Statistics Report;[13] full source breakdown for the other components is provided in Appendix: Reference Tables.

| Risk Group | Index | Population | Incidence (per 100k) | Composition | Source |
|---|---|---|---|---|---|
| PLWH | *i=1* | 1·2 M | 34·9 | Persons living with HIV (diagnosed + undiagnosed), 81% U.S.-born, 19% non-U.S.-born | CDC HIV Surv. 2024 |
| Medical comorbidities[a] | *i=2* | 40·0 M | 9·4 | Diabetes (38·4M), ESRD (808K), TNF-$\alpha$ inhibitors (1·5M), solid organ transplant (120K), other immunosuppression incl. chronic corticosteroids (500K); net of ∼890K overlap | CDC NDSR 2024; NIDDK 2023; OPTN/SRTR 2023 |
| Non-U.S.-born | *i=3* | 40·6 M | 11·7 | Total non-U.S.-born (47·8M) minus those in PLWH (227K) and Medical comorbidities (7·0M) groups | ACS 2023; Kerani 2020; Choi 2022 |
| U.S.-born | *i=4* | 253·4 M | 0·6 | Total U.S.-born (287·4M) minus those in PLWH (973K) and Medical comorbidities (33·0M) groups | Census 2023; derived |
| **Total** | | **335·2 M** | **3·1** | | |

Table S2 and Table S3 provide reference information for model parameters and sampling distributions used throughout the analysis.

**Table S2. Complete model parameters with sensitivity ranges and distributional assumptions.** [a]Stratum-specific ranges: $\epsilon_{s,HIV}$: 0·0041–0·013 (Ekramnia 95% UI)[9]; $\epsilon_{s,Med}$: 0·00042–0·00166 (±60%); $\epsilon_{s,NUSB}$: 0·00058–0·00097 (Ekramnia 95% UI); $\epsilon_{s,USB}$: 0·00029–0·0020 (wide exploratory). One-way sensitivity varies each parameter individually with full recalibration of $\beta$ and *Mtb* infection prevalences. LHS samples all 12 parameters simultaneously from the ranges shown (Table S3).

| Parameter | Symbol | PLWH | Medical | Non-U.S.-born | U.S.-born | Range | Source |
|---|---|---|---|---|---|---|---|
| Fast progression | $\varepsilon_f$ | 0·475 | 0·238 | 0·190 | 0·190 | ±50% | Derived (Fast Progression Rate [illegible] |
| Slow reactivation | $\varepsilon_s$ | 0·0069 | 0·00104 | 0·00074 | 0·00068 | See note[a] | Ekramnia et al. 2024[9] [illegible] |
| $L_f \rightarrow L_s$ transition | $\nu_f^{trans}$ | 0·30 | 0·30 | 0·30 | 0·30 | Fixed | Ferebee 1962[2] |
| $L_f$ clearance | $\nu_f^{clear}$ | *1.4×10^-6* | *1.4×10^-6* | *1.4×10^-6* | *1.4×10^-6* | Fixed | Negligible |
| $L_s$ clearance | $\nu_s^{clear}$ | 0·002 | 0·002 | 0·002 | 0·002 | 0·0005–0·005 | Vynnycky & Fine 1997 [illegible] |
| Fraction to $L_f$ | $p$ | 0·25 | 0·20 | 0·15 | 0·15 | Fixed | Vynnycky & Fine 1997 [illegible] |
| Transmission rate | $\beta$ | 5·70 | 5·70 | 5·70 | 5·70 | Recalibrated | Calibrated (Model Calibra [illegible] |
| Assortative mixing | $\varepsilon_i^{mix}$ | 0 | 0 | 0·70 | 0·70 | 0·40–0·90 | Menzies et al. 2018[4] |
| Reinfection protection | $\sigma$ | 0·79 | 0·79 | 0·79 | 0·79 | 0·50–1·00 | Andrews et al. 2012[14] |
| Treatment rate | $\gamma$ | 2·0 | 2·0 | 2·0 | 2·0 | 1·50–2·50 | CDC/WHO 2020[15] |
| Relapse rate | $\rho$ | 0·015 | 0·007 | 0·005 | 0·005 | Fixed | Romanowski et al. 2019 [illegible] |
| Stabilization | $\alpha_{stab}$ | 0·20 | 0·20 | 0·20 | 0·20 | 0·10–0·40 | Millet 2013,[16] Dobler 20 [illegible] |
| Population | $N$ | 1·2 M | 40·0 M | 40·6 M | 253·4 M | Fixed | Table 1 sources |
| Background mortality | $\mu$ | 0·012 | 0·025 | 0·009 | 0·010 | Fixed | CDC NVSS; USRDS 20 [illegible] |
| TB-specific mortality | $\mu_{TB}$ | 0·25 | 0·10 | 0·05 | 0·05 | Fixed | Corbett 2003,[17] Tiemersma [illegible] |
| Immigration rate | $\iota$ | 0 | 0 | 0·023 | 0 | Fixed | DHS 2023[1] |
| Efficacy | VE | 50% | 50% | 50% | 50% | 30–70% (P), 50–90% (O) | Tait et al. 2019[19] |
| Waning rate | $\omega$ | 0·10 (10-yr) | 0·10 (10-yr) | 0·10 (10-yr) | 0·10 (10-yr) | 5–15 yr | Modeling assumption [illegible] |
| Coverage rate | $\psi$ | 5%/yr | 5%/yr | 5%/yr | 5%/yr | 1–20% | Modeling assumption [illegible] |

**Table S3. LHS parameter sampling distributions (12 parameters, 500 draws).** Duration is sampled uniformly (5–15 yr) and converted to $\omega = 1/duration$. VE is sampled symmetrically around each scenario's central assumption (0·50 plausible, 0·70 optimistic), giving ranges of 0·30–0·70 and 0·50–0·90 respectively. Both ranges have the same ±20pp width to make the two scenarios' uncertainty intervals comparable. For each LHS draw, $\beta$ and stratum-specific *Mtb* infection prevalences are fully recalibrated to maintain TB incidence targets before evaluating vaccination impact.

| Parameter | Baseline | Range | Source / Rationale |
|---|---|---|---|
| VE (vaccine efficacy) | 0·50 / 0·70 | 0·30–0·70 / 0·50–0·90 | M72/AS01$_E$ trial 95% CI[19]; sampled symmetrically around each scenario's central VE |
| Duration (years) | 10 | 5–15 | Modeling assumption; symmetric around baseline |
| $\varepsilon_s$_HIV | 0·0069 | 0·0041–0·013 | Ekramnia et al. 2024 95% UI[9] |
| $\varepsilon_s$_Med | 0·00104 | 0·00042–0·00166 | ±60% composite |
| $\varepsilon_s$_NUSB | 0·00074 | 0·00058–0·00097 | Ekramnia et al. 2024 95% UI[9] |
| $\varepsilon_s$_USB | 0·00068 | 0·00029–0·0020 | Wide exploratory |
| $\varepsilon_f$ (base) | 0·19 | 0·12–0·28 | ±50%; derived from Vynnycky & Fine 1997[3] |
| $\nu_s^{clear}$ | 0·002 | 0·0005–0·005 | Vynnycky & Fine 1997[3] |
| $\sigma$ (reinfection) | 0·79 | 0·50–1·00 | Andrews et al. 2012[14] |
| $\varepsilon_i^{mix}$ (assortative) | 0·70 | 0·40–0·90 | Menzies et al. 2018[4] |
| $\gamma$ (treatment) | 2·0 | 1·50–2·50 | CDC/WHO 2020[15]; 5–8 month treatment |
| $\alpha_{stab}$ (stabilization) | 0·20 | 0·10–0·40 | Millet 2013[16]; 2·5–10 yr to stable *Mtb* infection |

### Direct and Indirect Vaccine Effects

Stratum-level results for the All-High-Risk strategy are presented in Figure S2. We decomposed total impact into direct and indirect components using a static force-of-infection method.[20–22] Clinical-only strategies (Medical comorbidities and PLWH + Medical) had the lowest indirect fractions, around 11% (Table S4).

We decomposed total impact into direct and indirect components using a static force-of-infection method.[20,22,23] We ran three scenarios at equilibrium: (1) no vaccination, (2) vaccination with the force of infection updated dynamically as TB prevalence changed, and (3) vaccination with the force of infection held fixed at baseline values. Scenario 3 isolates the direct effect of vaccination (reduced progression in vaccinated individuals) because $\lambda_i$ does not change even as infectious prevalence falls. The indirect effect (reduced transmission benefiting unvaccinated individuals) is the difference between scenarios 2 and 3.

Clinical-only strategies (Medical comorbidities and PLWH + Medical) had the lowest indirect fractions (around 11%).

**Table S4. Direct and indirect effects of vaccination by strategy under optimistic (VE = 70%, vaccination rate = 50%/yr) and plausible (VE = 50%, strategy-specific vaccination rate) scenarios with 10-year protection.** Total Prevented = total cases prevented per year. Direct = cases prevented through reduced progression in vaccinated individuals. Indirect = cases prevented through reduced transmission benefiting unvaccinated individuals. Plausible scenario uses VE = 50% with strategy-specific vaccination rates (5%/yr for most strategies, 10%/yr for PLWH, 2%/yr for All *Mtb*-Infected). Optimistic scenario uses VE = 70% with vaccination rate = 50%/yr for all strategies. Both scenarios use 10-year duration of protection. Varying assortative mixing ($\varepsilon_{mix}$) from 0 to 0·90 changes total cases prevented by $<50$, so the direct/indirect split is not sensitive to mixing assumptions. PLWH = persons living with HIV.

| Strategy | Scenario | Total Prevented | Direct | Indirect | % Direct | % Indirect |
|---|---|---|---|---|---|---|
| All-High-Risk | Plausible | 1,342 | 1,157 | 185 | 86·2% | 13·8% |
| PLWH + Non-U.S.-Born | Plausible | 921 | 770 | 150 | 83·7% | 16·3% |
| All *Mtb*-Infected | Plausible | 727 | 614 | 113 | 84·5% | 15·5% |
| PLWH + Medical | Plausible | 623 | 555 | 68 | 89·0% | 11·0% |
| Medical comorbidities | Plausible | 559 | 496 | 63 | 88·7% | 11·3% |
| PLWH | Plausible | 110 | 85 | 25 | 77·0% | 23·0% |
| All-High-Risk | Optimistic | 5,380 | 4,774 | 606 | 88·7% | 11·3% |
| PLWH + Non-U.S.-Born | Optimistic | 4,325 | 3,780 | 545 | 87·4% | 12·6% |
| All *Mtb*-Infected | Optimistic | 6,005 | 5,403 | 602 | 90·0% | 10·0% |
| PLWH + Medical | Optimistic | 2,466 | 2,253 | 212 | 91·4% | 8·6% |
| Medical comorbidities | Optimistic | 2,215 | 2,021 | 194 | 91·2% | 8·8% |
| PLWH | Optimistic | 255 | 216 | 39 | 84·7% | 15·3% |

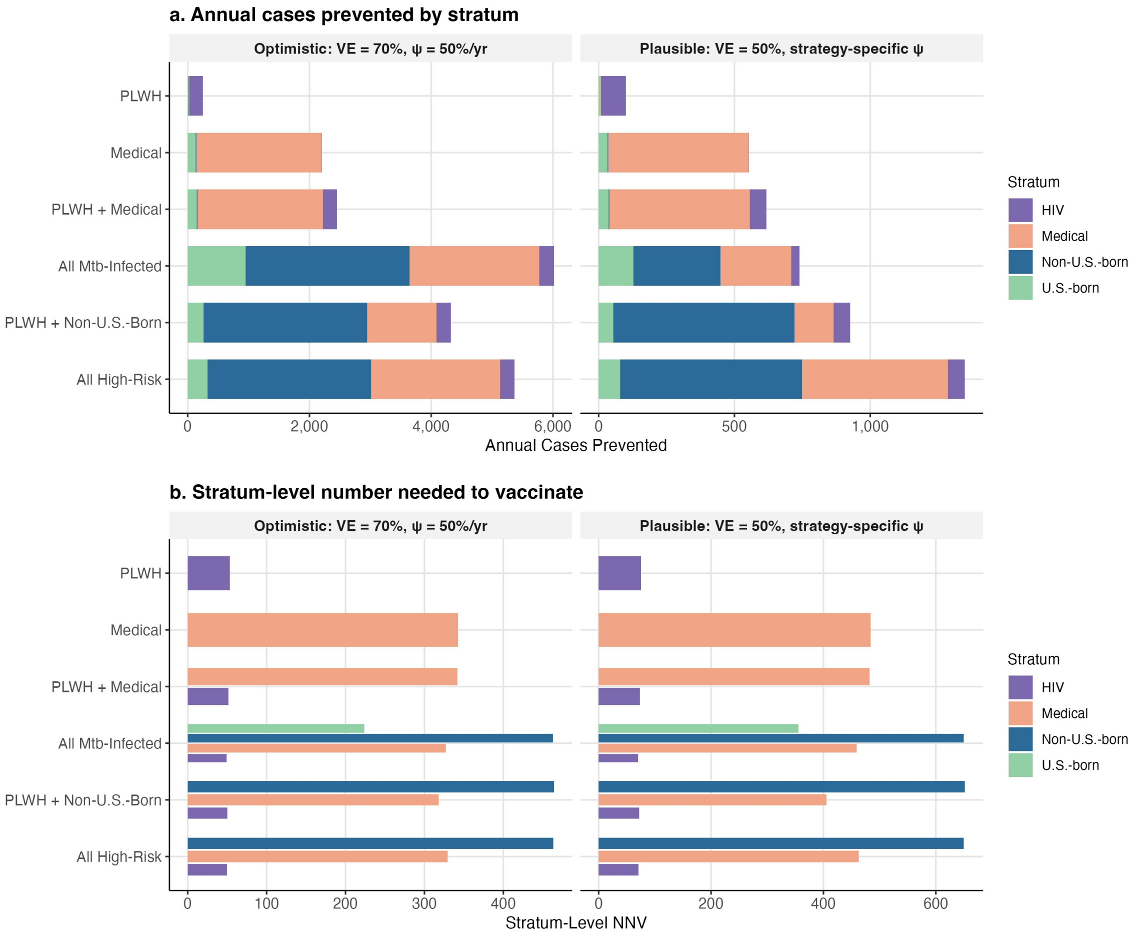

**Figure S2. Stratum-level vaccination impact by strategy under both scenarios.** a: Annual cases prevented by stratum (stacked). b: Stratum-level NNV for vaccinated strata only (dodged). Left panels: optimistic scenario (VE = 70%, vaccination rate = 50%/yr for all strategies). Right panels: plausible scenario (VE = 50%, strategy-specific vaccination rate per Table 1). Non-targeted strata (for example, U.S.-born under All-High-Risk) receive cases prevented entirely from indirect transmission effects. Duration of protection = 10 years in both scenarios. PLWH = persons living with HIV. NNV = number needed to vaccinate.

### Sensitivity and Uncertainty Analysis

#### One-Way Sensitivity

Strategy rankings were unchanged under one-at-a-time variation of all 11 natural history parameters (Figure S3).

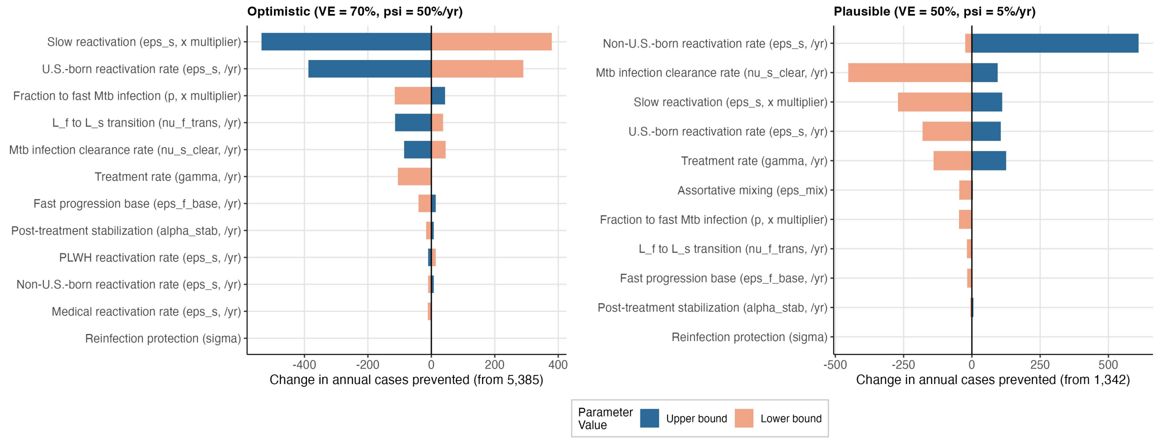

**Figure S3. One-way sensitivity analysis, All-High-Risk strategy, under optimistic (VE = 70%, vaccination rate ($\psi$) = 50%/yr, left) and plausible (VE = 50%, vaccination rate ($\psi$) = 5%/yr, right) scenarios with 10-year protection.** Parameters ranked by range of change in annual cases prevented from each scenario's baseline (5,380 optimistic; 1,342 plausible). Full recalibration of $\beta$ and *Mtb* infection prevalences at each parameter value.

#### Latin Hypercube Sampling and PRCC

Uncertainty intervals from 500 LHS draws under each scenario are reported alongside point estimates in Table 2. PRCC analysis identified vaccine efficacy and the waning rate $\omega$ as the dominant drivers of cases prevented (Figure S4).

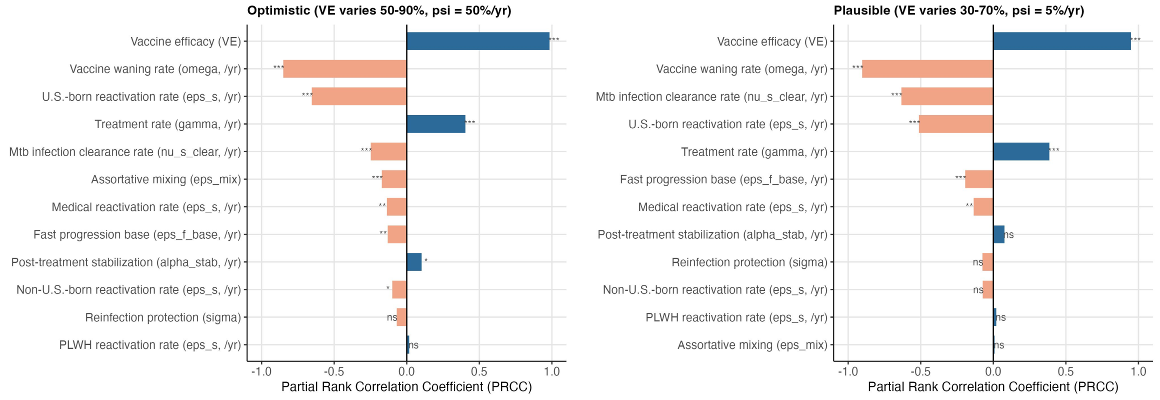

**Figure S4. Partial Rank Correlation Coefficients (PRCC) for cases prevented, All-High-Risk strategy, under optimistic (vaccination rate ($\psi$) = 50%/yr, left) and plausible (vaccination rate ($\psi$) = 5%/yr, right) scenarios.** VE varies 50–90% in the optimistic scenario and 30–70% in the plausible scenario, sampled symmetrically around each scenario's central VE assumption. Parameters ranked by PRCC magnitude, controlling for all other parameters. Vaccine efficacy and the waning rate $\omega$ are the dominant drivers in both scenarios.

### Vaccination Coverage-Impact Analysis

NNV was essentially constant across coverage rates from 1% to 50% per year for every strategy (e.g., All-High-Risk: 513–517 at VE = 50%), and cases prevented scaled linearly with coverage (Table S5). This linear scaling reflects the reactivation-dominated nature of the U.S. epidemic: each vaccinated person faces an independent probability of progressing to TB disease, so per-person benefit does not decline as coverage expands.

Effective coverage (the fraction of *Mtb*-infected individuals in vaccinated compartments at equilibrium) was lower than the annual vaccination rate because vaccine protection wanes over time (Table S6). At vaccinate rate =5% per year, effective coverage ranged from 0·2% (PLWH) to 27·9% (All *Mtb*-Infected). Even at vaccination rate =20% per year, no strategy exceeded 61% effective coverage.

**Table S5. Coverage-impact analysis for all strategies (VE = 50%, 10-year protection, $\psi$ = 1–50%/yr).**

| Strategy | Vaccination rate $\psi$ (%/yr) | Cases Prevented | % Reduction | Annual Vaccinations | NNV | Effective Coverage (%) |
|---|---|---|---|---|---|---|
| All-High-Risk | 1% | 348 | 3·4% | 178,198 | 513 | 6·2 |
| | 5% | 1,342 | 12·9% | 693,688 | 517 | 24·2 |
| | 10% | 2,108 | 20·3% | 1,084,980 | 515 | 37·9 |
| | 20% | 2,944 | 28·3% | 1,511,540 | 513 | 53·0 |
| | 30% | 3,393 | 32·7% | 1,739,949 | 513 | 61·8 |
| | 50% | 3,867 | 37·2% | 1,979,303 | 512 | 70·4 |
| PLWH + Non-U.S.-Born | 1% | 234 | 2·3% | 125,552 | 536 | 4·5 |
| | 5% | 921 | 8·9% | 500,875 | 544 | 17·7 |
| | 10% | 1,486 | 14·3% | 803,783 | 541 | 28·6 |
| | 20% | 2,174 | 20·9% | 1,165,326 | 536 | 41·8 |
| | 30% | 2,571 | 24·7% | 1,378,822 | 536 | 49·5 |
| | 50% | 3,001 | 28·9% | 1,608,771 | 536 | 57·7 |
| All *Mtb*-Infected | 1% | 392 | 3·8% | 201,359 | 513 | 7·2 |
| | 5% | 1,515 | 14·6% | 780,135 | 515 | 27·9 |
| | 10% | 2,375 | 22·9% | 1,215,417 | 512 | 43·6 |
| | 20% | 3,310 | 31·9% | 1,685,837 | 509 | 60·7 |
| | 30% | 3,808 | 36·7% | 1,934,990 | 508 | 69·7 |
| | 50% | 4,334 | 41·7% | 2,198,988 | 507 | 78·9 |
| PLWH + Medical | 1% | 166 | 1·6% | 66,287 | 400 | 2·4 |
| | 5% | 623 | 6·0% | 256,555 | 412 | 9·2 |
| | 10% | 972 | 9·4% | 399,480 | 411 | 14·4 |
| | 20% | 1,351 | 13·0% | 553,937 | 410 | 19·9 |
| | 30% | 1,555 | 15·0% | 637,259 | 410 | 22·9 |
| | 50% | 1,775 | 17·1% | 726,897 | 410 | 26·2 |
| Medical comorbidities | 1% | 149 | 1·4% | 65,118 | 437 | 2·3 |
| | 5% | 559 | 5·4% | 252,112 | 451 | 9·0 |
| | 10% | 872 | 8·4% | 392,663 | 450 | 14·1 |
| | 20% | 1,213 | 11·7% | 544,633 | 449 | 19·6 |
| | 30% | 1,396 | 13·4% | 626,416 | 449 | 22·6 |
| | 50% | 1,593 | 15·3% | 714,178 | 448 | 25·8 |
| PLWH | 1% | 26 | 0·3% | 1,171 | 45 | 0·0 |
| | 5% | 74 | 0·7% | 4,474 | 60 | 0·2 |
| | 10% | 110 | 1·1% | 6,898 | 63 | 0·3 |
| | 20% | 148 | 1·4% | 9,468 | 64 | 0·4 |
| | 30% | 164 | 1·6% | 10,564 | 64 | 0·4 |
| | 50% | 180 | 1·7% | 11,600 | 64 | 0·4 |

$\psi$ = annual vaccination rate (1–50%/yr). Cases Prevented = cases prevented (annual reduction relative to unvaccinated baseline). % Reduction = percent reduction in overall TB burden. Annual Vaccinations = annual vaccinations. NNV = number needed to vaccinate (annual vaccinations ÷ annual cases prevented). Effective Coverage = effective coverage (% of *Mtb*-infected individuals in vaccinated compartments at equilibrium, overall population). PLWH = persons living with HIV. VE = 50%, 10-year protection. In this analysis, annual vaccination rate ($\psi$) is varied uniformly for each strategy. Values at $\psi = 5\%$ may differ slightly from the main results table, which uses strategy-specific vaccination rates. PLWH uses $\psi = 10\%$ and All *Mtb*-Infected uses $\psi = 2\%$ in the primary analysis.

**Table S6. Effective coverage at equilibrium by strategy and population stratum under optimistic (VE = 70%, vaccination rate = 50%/yr) and plausible (VE = 50%, strategy-specific vaccination rate) scenarios with 10-year protection.** Effective coverage is the percentage of *Mtb*-infected individuals in vaccinated compartments at equilibrium. Stratum-specific = effective coverage within each individual stratum. Targeted Strata = pooled effective coverage across all strata receiving vaccination. Overall = effective coverage across the full national *Mtb* infection reservoir (21·7 million persons), including untargeted strata. Plausible scenario uses VE = 50% with strategy-specific vaccination rates (Table 1). Optimistic scenario uses VE = 70% with annual vaccination rate = 50%/yr for all strategies. Both scenarios use 10-year duration of protection. "— indicates the stratum is not targeted under that strategy. Effective coverage is lower than the vaccination rate because vaccine protection wanes over time (vaccination waning rate = 0·10/yr). PLWH = persons living with HIV.

| Strategy | Scenario | Stratum-specific (%) | | | | Targeted strata (%) | Overall (%) |
|---|---|---|---|---|---|---|---|
| | | PLWH | Medical | Non-U.S.-born | U.S.-born | | |
| All-High-Risk | Plausible | 29·9 | 28·1 | 27·1 | — | 27·5 | 24·3 |
| PLWH + Non-U.S.-Born | Plausible | 29·9 | 6·5 | 27·1 | — | 19·6 | 17·3 |
| All *Mtb*-Infected | Plausible | 14·4 | 13·5 | 12·9 | 14·4 | 13·2 | 13·2 |
| PLWH + Medical | Plausible | 29·8 | 28·1 | — | — | 28·2 | 9·3 |
| Medical comorbidities | Plausible | — | 28·1 | — | — | 28·1 | 9·1 |
| PLWH | Plausible | 46·1 | — | — | — | 46·1 | 0·3 |
| All-High-Risk | Optimistic | 81·3 | 79·8 | 78·9 | — | 79·3 | 70·6 |
| PLWH + Non-U.S.-Born | Optimistic | 81·3 | 41·5 | 78·9 | — | 65·2 | 57·9 |
| All *Mtb*-Infected | Optimistic | 81·3 | 79·8 | 78·9 | 81·5 | 79·5 | 79·5 |
| PLWH + Medical | Optimistic | 81·3 | 79·8 | — | — | 79·8 | 26·4 |
| Medical comorbidities | Optimistic | — | 79·8 | — | — | 79·8 | 25·9 |
| PLWH | Optimistic | 81·3 | — | — | — | 81·3 | 0·5 |

### Time-to-Impact Analysis

We integrated the ODE system forward from baseline equilibrium to track how vaccination impact accumulates over time (Table S7). Cumulative NNV (cumulative vaccinations divided by cumulative cases prevented) was highest in the early program years and decreased as annual impact approached equilibrium (Table S8).

**Table S7. Time-to-impact trajectories: annual cases prevented and cumulative impact by strategy under optimistic (VE = 70%, annual vaccination rate = 50%/yr) and plausible (VE = 50%, strategy-specific annual vaccination rate) scenarios with 10-year protection.** Annual Cases Prevented = reduction in annual cases relative to the unvaccinated baseline at the specified program year. Cumulative Prevented = total cases prevented from program initiation through the specified year. Yr = program year after vaccination initiation. Plausible scenario uses VE = 50% with strategy-specific vaccination rates (5%/yr for most strategies, 10%/yr for PLWH, 2%/yr for All *Mtb*-Infected). Optimistic scenario uses VE = 70% with vaccination rate = 50%/yr for all strategies. Both scenarios use 10-year duration of protection. Strategy rankings are preserved at every time horizon under both scenarios. PLWH = persons living with HIV.

| Strategy | Scenario | Annual cases prevented | | | | Cumulative cases prevented | | | |
|---|---|---|---|---|---|---|---|---|---|
| | | Yr 1 | Yr 5 | Yr 10 | Yr 30 | Yr 1 | Yr 5 | Yr 10 | Yr 30 |
| All-High-Risk | Plausible | 115 | 701 | 1,070 | 1,334 | 115 | 1,714 | 6,142 | 31,354 |
| PLWH + Non-U.S.-Born | Plausible | 75 | 465 | 719 | 912 | 75 | 1,130 | 4,091 | 21,218 |
| All *Mtb*-Infected | Plausible | 51 | 330 | 530 | 718 | 51 | 790 | 2,939 | 16,120 |
| PLWH + Medical | Plausible | 55 | 327 | 496 | 615 | 55 | 805 | 2,863 | 14,511 |
| Medical comorbidities | Plausible | 49 | 293 | 444 | 550 | 49 | 721 | 2,564 | 12,983 |
| PLWH | Plausible | 12 | 62 | 88 | 101 | 12 | 157 | 532 | 2,497 |
| All-High-Risk | Optimistic | 1,376 | 4,812 | 5,300 | 5,380 | 1,376 | 14,228 | 39,508 | 146,724 |
| PLWH + Non-U.S.-Born | Optimistic | 911 | 3,498 | 4,104 | 4,325 | 911 | 9,980 | 28,983 | 114,360 |
| All *Mtb*-Infected | Optimistic | 1,503 | 5,330 | 5,885 | 6,005 | 1,503 | 15,697 | 43,734 | 163,091 |
| PLWH + Medical | Optimistic | 657 | 2,240 | 2,447 | 2,466 | 657 | 6,679 | 18,397 | 67,689 |
| Medical comorbidities | Optimistic | 587 | 2,009 | 2,195 | 2,215 | 587 | 5,986 | 16,496 | 60,748 |
| PLWH | Optimistic | 70 | 232 | 254 | 255 | 70 | 696 | 1,912 | 7,021 |

**Table S8. Cumulative number needed to vaccinate (NNV) by strategy and program year under optimistic (VE = 70%, vaccination rate = 50%/yr) and plausible (VE = 50%, strategy-specific annual vaccination rate) scenarios with 10-year protection.** Cumulative NNV = cumulative vaccinations ÷ cumulative cases prevented through the specified program year. Values decrease over time as annual impact approaches equilibrium. Annual Equilibrium = NNV at steady state (50-year integration) from Table 2. Yr = program year after vaccination initiation. Plausible scenario uses VE = 50% with strategy-specific annual vaccination rates. Optimistic scenario uses VE = 70% with vaccination rate = 50%/yr for all strategies. Both scenarios use 10-year duration of protection. PLWH converges fastest due to its small, high-incidence target population. NNV = number needed to vaccinate. PLWH = persons living with HIV.

| Strategy | Scenario | Cumulative NNV at program year | | | | | | | Annual equilibrium |
|---|---|---|---|---|---|---|---|---|---|
| | | Yr 1 | Yr 2 | Yr 3 | Yr 5 | Yr 10 | Yr 20 | Yr 30 | |
| All-High-Risk | Plausible | 8,122 | 4,548 | 3,163 | 2,025 | 1,200 | 801 | 687 | 510 |
| PLWH + Non-U.S.-Born | Plausible | 8,723 | 4,881 | 3,390 | 2,164 | 1,277 | 850 | 727 | 537 |
| All *Mtb*-Infected | Plausible | 8,380 | 4,709 | 3,280 | 2,100 | 1,238 | 820 | 697 | 508 |
| PLWH + Medical | Plausible | 6,376 | 3,595 | 2,509 | 1,616 | 963 | 644 | 552 | 413 |
| Medical comorbidities | Plausible | 7,026 | 3,945 | 2,752 | 1,772 | 1,056 | 707 | 607 | 451 |
| PLWH | Plausible | 1,012 | 598 | 416 | 266 | 158 | 105 | 91 | 68 |
| All-High-Risk | Optimistic | 4,475 | 2,424 | 1,607 | 1,013 | 641 | 487 | 443 | 366 |
| PLWH + Non-U.S.-Born | Optimistic | 4,925 | 2,675 | 1,783 | 1,127 | 704 | 520 | 468 | 378 |
| All *Mtb*-Infected | Optimistic | 4,555 | 2,457 | 1,623 | 1,019 | 641 | 485 | 441 | 363 |
| PLWH + Medical | Optimistic | 3,514 | 1,912 | 1,271 | 804 | 509 | 387 | 353 | 292 |
| Medical comorbidities | Optimistic | 3,860 | 2,099 | 1,394 | 881 | 558 | 425 | 387 | 320 |
| PLWH | Optimistic | 599 | 328 | 218 | 137 | 86 | 64 | 58 | 48 |

### NNV Benchmarking

To put our NNV estimates in context, we compared them with NNV values from published TB vaccine modeling studies and from other recommended adult vaccines (Figure S5). There is no universal NNV threshold that defines an acceptable vaccination program,[24] but benchmarks from other vaccines and TB models provide useful reference points. Direct comparisons across vaccines are limited by differences in disease burden, outcome severity, target population risk, and metric definition. This study reports annual equilibrium NNV, whereas Harris et al. report cumulative NNV over the 2025–2050 horizon, and the comparator vaccines use a mix of single-year and multi-year metrics.

Harris et al. modeled age-targeted TB vaccination in China and reported cumulative NNV per case averted (2025–2050) for older adults (aged 60–64) at 60% VE and 10-year protection.[21] For post-infection vaccines effective in latency and recovered individuals (L&R), the NNV was 292 (UI: 257–365), while the broadest vaccine profile (effective pre- and post-infection) had an NNV of 230 (UI: 199–269). Pre-infection vaccines had higher NNV (1,022, UI: 752–1,318), which fits with our finding that vaccinating persons with existing *Mtb* infection is more efficient in reactivation-dominated settings. Our estimates (68–537 across primary strategies at 50% VE) overlap the Harris post-infection range despite differences in setting (U.S. vs. China), VE assumption (50% vs. 60%), and model structure (risk-stratified vs. age-stratified). To our knowledge, Harris et al. is the only published TB vaccine modeling study that reports NNV directly. Other studies modeling TB vaccination in the US,[25] low-incidence settings,[26] and high-burden countries[20,27,28] have reported other outcomes including percentage reductions in incidence, cumulative cases averted, and cost-effectiveness.

Our NNV for vaccinating all high-risk groups (510, 95% UI: 284–1,631 under the plausible scenario) is also comparable to NNV values for other recommended adult vaccination programs. The NNV for PCV-13 vaccination against community-acquired pneumonia (CAP) in adults 65 years was 234–576 for all CAP over 5 years and 1,620 for hospitalized CAP over 1 year,[29,30] and influenza vaccination in older adults had an NNV of 777 to prevent one hospitalization.[31] The NNV for vaccinating only PLWH (68, 95% UI: 19–1,674 under the plausible scenario) is among the lowest for any adult vaccination program targeting a serious disease outcome.

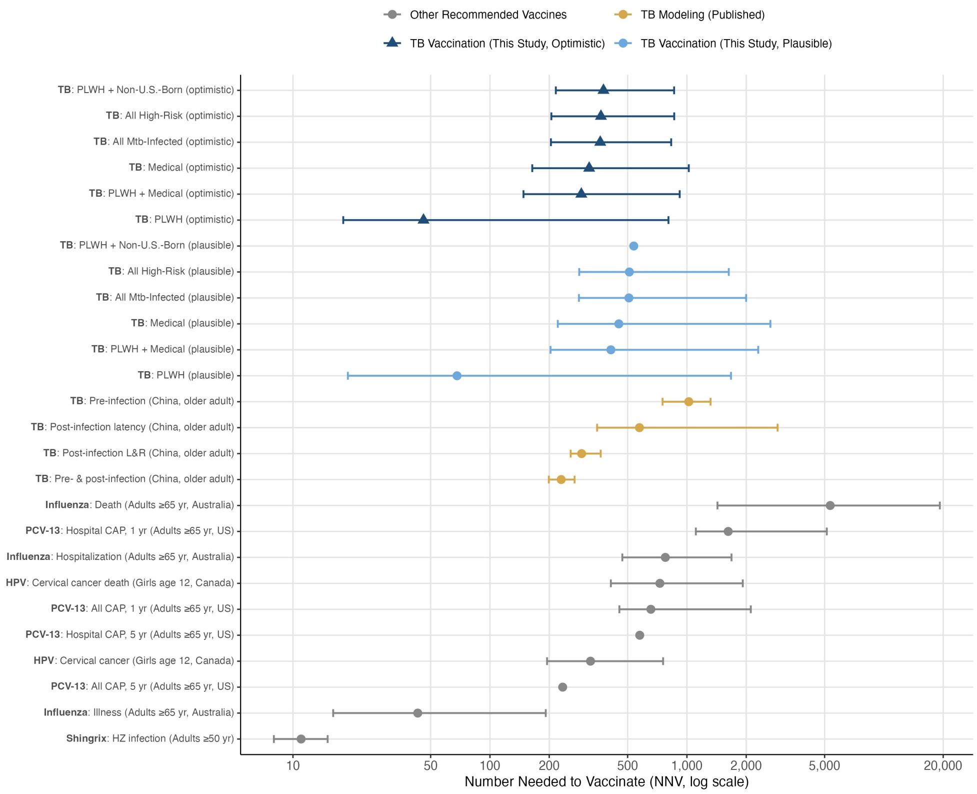

**Figure S5. NNV benchmarking against published TB vaccine modeling estimates and established vaccines.** Model-estimated NNV for each strategy under the optimistic scenario (dark blue triangles, VE = 70%, vaccination rate = 50%/yr, 10-year protection) and the plausible scenario (light blue circles, VE = 50%, strategy-specific annual vaccination rate, 10-year protection) compared with published TB vaccine NNV from Harris et al. 2019 (gold, VE = 60%, 10-year protection, 70% coverage, older adults aged 60–64 in China)[23] and NNV values for other recommended adult vaccines (gray). Under the optimistic scenario, NNV ranges from 48 (PLWH) to 378 (PLWH + Non-U.S.-Born), with All-High-Risk at 366 and All *Mtb*-Infected at 363. Error bars show 95% uncertainty intervals from Latin Hypercube Sampling (500 draws per scenario, varying 12 parameters with full recalibration at each draw, for this study's TB points), uncertainty intervals (Harris 2019), or 95% confidence intervals (established vaccines). Vaccine profiles in the Harris estimates are pre-infection (effective before *Mtb* infection only), post-infection latency (effective in latent *Mtb* infection only), post-infection L&R (effective in latency or after recovery from active disease), and pre- & post-infection (effective regardless of infection status). Sources for comparator vaccines are Shingrix[32], influenza[31], PCV-13[29,30], and HPV.[33] CAP = community-acquired pneumonia. L&R = latency or recovered from active disease.

[8] Centers for Disease Control and Prevention. Reported tuberculosis in the United States, 2024: National tuberculosis surveillance system. Technical report, U.S. Department of Health and Human Services, CDC, Atlanta, GA, 2024. 10,388 reported TB cases; calibration target.

[9] Mina Ekramnia, Yunfei Li, Maryam B. Haddad, Suzanne M. Marks, J. Steve Kammerer, Nicole A. Swartwood, Ted Cohen, Jeffrey W. Miller, C. Robert Horsburgh, Joshua A. Salomon, and Nicolas A. Menzies. Estimated rates of progression to tuberculosis disease for persons infected with Mycobacterium tuberculosis in the United States. *Epidemiology*, 35(2):164–173, 2024. doi: 10.1097/EDE.0000000000001707.

[10] Daniel E. Jonas, Sheila Riley, Leila Lee, et al. *Screening for Latent Tuberculosis Infection in Adults: An Evidence Review for the U.S. Preventive Services Task Force*. Agency for Healthcare Research and Quality (US), 2023. URL `http://www.ncbi.nlm.nih.gov/books/NBK591768/`. Accessed July 3, 2024.

[11] K. Romanowski, B. Baumann, C. A. Basham, F. Ahmad Khan, G. J. Fox, and J. C. Johnston. Long-term all-cause mortality in people treated for tuberculosis: A systematic review and meta-analysis. *Lancet Infectious Diseases*, 19(10):1129–1137, 2019.

[12] C. C. Dobler, A. B. J. Crawford, P. J. Jelfs, G. L. Gilbert, and G. B. Marks. Recurrence of tuberculosis in a low-incidence setting. *European Respiratory Journal*, 33(1):160–167, 2009.

[13] Centers for Disease Control and Prevention. National diabetes statistics report: Estimates of diabetes and its burden in the United States. Technical report, U.S. Department of Health and Human Services, Atlanta, GA, 2024. 2021 data: 38.4M total diabetes (11.6% of US population).

[14] Jason R. Andrews, Farzad Noubary, Rochelle P. Walensky, Rodrigo Cerda, Elena Losina, and C. Robert Horsburgh. Risk of progression to active tuberculosis following reinfection with *Mycobacterium tuberculosis*. *Clinical Infectious Diseases*, 54(6):784–791, 2012. doi: 10.1093/cid/cir951.

[15] Centers for Disease Control and Prevention. Updated guidelines for testing for latent tuberculosis infection. Technical report, U.S. Department of Health and Human Services, 2020. Recommends LTBI screening for PLWH and other immunocompromised populations.
